# Specific Lipid Abnormalities Are Inherently Associated with Late-Onset Alzheimer’s Disease

**DOI:** 10.1101/2025.08.08.25333304

**Authors:** Bruce M. Cohen, Eunjung Koh, Kandice R. Levental, Ilya Levental, Kai-Christian Sonntag

## Abstract

**INTRODUCTION:** Lipid abnormalities have been observed in brain, CSF, and blood in association with late-onset Alzheimer’s disease (LOAD). It is unknown which abnormalities are precursors to LOAD and which are concomitants of illness or its treatment. Inherent abnormalities can be identified in induced pluripotent stem cell (iPSC)-derived neural lines.

**METHODS:** iPSC lines of patients with LOAD or healthy individuals were differentiated to astrocytes. Lipidomics analyses were performed on whole cell and mitochondrial extracts.

**RESULTS:** Large reductions in cholesterol esters (CE) and imbalances in fatty acids (FA) were observed in LOAD-associated cells or their mitochondria. There were only modest differences in other lipid classes, including membrane structural lipids.

**DISCUSSION:** The findings identify abnormalities in CE and FA as likely precursors to LOAD. These differences implicate mechanisms contributing to disease pathogenesis. Further study may lead to early interventions to prevent or delay LOAD.

## BACKGROUND

Over 90% of dementias present after the age of 65, and in most of these cases, late-onset Alzheimer’s disease (LOAD) is diagnosed.^1^

Unlike early-onset Alzheimer’s disease (EOAD), which is usually the consequence of highly penetrant variants in single genes, the inherent risk for LOAD is determined by interactions among numerous, possibly hundreds, of genes with low penetrance.^2^ These genes underlie multiple processes needed for cell development, repair and remodeling, and responses to aging and environmental exposures. Through effects on all these functions, they determine risk for LOAD.

Among the inherited factors linked to LOAD, both single gene findings and pathway analyses suggest that differences in lipid metabolism strongly affect risk.^2–4^ This includes gene sets associated with cholesterol synthesis, transport, and metabolism, as well as gene sets associated with fatty acid (FA) and phospholipid synthesis and metabolism.^2,5, 6^

The strongest gene association for LOAD is with *APOE*, and APOE4 homozygosity increases risk over 10-fold.^7 8^ *APOE* codes for a lipoprotein that transports lipids, predominantly cholesterol and cholesterol esters, as well as FA, between and within cells.^8^ APOE4 may increase the risk for LOAD both through its effects on lipid transport in brain, and through its effects on lipid accumulations on the walls of blood vessels supplying the brain.^9, 10^

Epidemiologic studies also suggest an association of lipids with LOAD. Specifically, diets rich in saturated fats can increase the risk for LOAD.^11^ And high levels of some fats and cholesterols in blood, packaged with APOE in low-density lipoproteins (LDL) are associated with an increased risk for dementia.^12^ Interventions that reduce cholesterol levels, including treatment with statins, reduce that risk.^13^

However, if there is an intact blood-brain barrier, plasma cholesterol does not freely enter the brain.^14, 15^ Rather, cholesterol is made in the brain, and there is evidence, from studies of cholesterol and its precursors in CSF, that patients with LOAD have decreased, not increased, production of cholesterol in brain.^15, 16^ The association of blood and dietary cholesterol with poorer brain health may largely be a consequence of the effects of blood cholesterols on vascular health.^17, 18^ Specifically, atherosclerosis is associated with decreased blood flow to brain, thereby compromising brain function and repair. In addition, damage to vasculature integrity, especially to the blood-brain barrier, exposes the brain to toxins in the circulation.

The role of FA is more debated than the role of cholesterols, but elevated saturated fats in diet and blood may be associated with an increased risk for dementia. In this case, as in the case of cholesterols, the causal mechanisms may be indirect. Saturated FA, versus unsaturated or polyunsaturated FA, induce inflammatory activity with widespread toxicity, including damage to the cardiovascular system and, through blood-borne inflammatory molecules, in the brain.^19^

It is easy to understand why lipid types and levels in brain might directly contribute to risk for LOAD. Most cellular processes occur across, along, or within membranous elements and organelles of cells.^20^ Lipids are key components of all cell membranes, and lipid content abnormalities can alter many physiologic functions known to be associated with LOAD. These include glucose uptake and glycolysis, protein processing in the Golgi apparatus and endoplasmic reticulum (ER), energy production in mitochondria, the digestion and elimination of toxic compounds in lysosomes and autophagosomes, the storage and release of metabolic precursors, and the storage and release of signaling molecules, which include Ca++, cholesterol esters, and FA that modulate cell functions.^20–22^ And A-beta (Aβ) amyloid fragment production, underlying the amyloid plaques often seen in LOAD, has been linked to the lipid composition of cell membranes.^13^

In this paper, we briefly review reports on lipid content observed in post-mortem brain, CSF, and plasma in people with LOAD. Some studies included people with minimal cognitive impairment (MCI), which often precedes LOAD. Because the measures in these studies were determined in samples from people who were symptomatic, it is not known with certainty which of the abnormalities observed are basic factors underlying risk for LOAD, and which are a consequence of illness and its treatment.^23^

To address this question, we studied lipids in reprogrammed cells obtained from individuals with LOAD. Fibroblasts or blood cells were converted to induced pluripotent stem cell lines (iPSC), differentiated into neural progenitor cells (NPCs), and then into more mature brain cells. Such reprogrammed cells, produced from iPSC lines, lack most or all epigenetic or other features associated with aging, illness, and its treatment. Lipid abnormalities in these cells are, therefore, inherent to the lines. They are likely to be abnormalities that underly neuropathological processes contributing to the development of LOAD.

### Past Studies of Lipid Classes in Individuals with LOAD

Most lipids analyzed in people with LOAD fall into the following classes: (1) cholesterol and cholesterol esters, (2) glycerophospholipids and their derivatives, and (3) sphingolipids, including ceramides and sphingomyelin.^20^ Cholesterol, glycerophospholipids, and sphingolipids are present at substantial concentrations in all cell membranes, but their relative proportions differ among membranes.^20, 23^ Cholesterol is highest in the external cell membrane, and phosphatidylcholine, one of several common glycerophospholipids, is the highest concentration lipid in most other (internal) cell membranes. Sphingolipids are present at lower concentrations in both external and internal membranes. Lipids in each class can have side chains, most commonly, FA. These can be exchanged among lipids of different classes. They are also seen free or within lipid droplets in cells.

### Cholesterol and its derivatives in LOAD

Cholesterols in most tissues, including brain, exist in 2 pools, neutral cholesterols and cholesterols esterified with FAs (CE). In membranes, neutral cholesterols are the core elements of lipid rafts that house membrane-associated proteins, including receptors, ion channels, and enzyme complexes, all necessary for basic cell function.^21^ Cholesterol is observed at especially high concentrations in the mitochondrial membranes that are essential sites for oxidative phosphorylation. Although elevated cholesterol in blood is associated with a higher risk for dementia, cholesterol does not appear to be increased in peripheral cell or brain cell membranes in people with LOAD.^24^ Cholesterol levels in brain increase with age, and while they are not substantially altered in post-mortem brain in LOAD, both cholesterol synthesis and degradation may be reduced in LOAD.^25^ These changes may be both a consequence and a driver of Aβ formation.^23^ That is, Aβ affects membrane composition and membrane composition affects amyloid processing.

CE are storage and signaling molecules. CE stored in lipid droplets within cells can be converted to cholesterol.^26^ As a messenger, CE can directly influence numerous cell processes including those related to bioenergetics, protein processing, autophagy, and inflammation.^26^ Lipid droplets, which also contain other neutral lipids, have been reported to be elevated in brain in post-mortem studies of LOAD.^8, 27^ CEs were observed to be elevated in frontal cortex^28^ or only in diseased entorhinal cortex and not frontal cortex in association with LOAD.^29^ It is not known if these increases are precursors or consequences of neurodegeneration in LOAD.^21^ *In vivo*, CE was observed to be decreased in plasma in people with LOAD.^30^ Since that first report, it has consistently been observed that CE is substantially decreased in both plasma and CSF of people with LOAD.^31, 32^ Low CE provided good classification accuracy for identifying individuals with LOAD.^32^ As a possible mechanism for these findings, cholesterol esterification rate was reduced in CSF from patients with LOAD versus healthy controls. ^33^ And genes associated with CE metabolism are also associated with LOAD.^32^ Addressing the stage at which this abnormality occurs, individuals with MCI have levels of CE between those seen in people with LOAD and healthy comparison subjects. In addition, people with MCI with lower CE are more likely to develop LOAD on follow-up.^24^ These genomic and mechanistic associations along with replicated observations of substantially low CEs *in vivo* in LOAD, and in people with MCI who proceed to LOAD, suggest that low CE might be a precursor of risk for LOAD.

### Glycerophospholipids and their derivatives in LOAD

Among glycerophospholipids, phosphatidylcholine (PC) has been most studied regarding an association with LOAD. This is in part because PC is the most prominent glycerophospholipid. More so, it is due to the observation that dysfunction, shrinkage, and death of long axon cholinergic neurons in brain are among the earliest and strongest signs of Alzheimer’s disease.^34, 35^ These neurons require choline both for use in PC as a structural lipid and for the production of the neurotransmitter acetylcholine. Brain choline uptake and metabolism decreases with age, and this decline may increase the risk for degeneration of cholinergic neurons.^36, 37^ Medications increasing cholinergic signaling were the first drugs developed and marketed for the treatment of LOAD and remain in use today.^38^

PC has most often been reported to be reduced in post-mortem tissue, as well as in CSF and blood, from individuals with LOAD.^3, 21^ However, some studies observed increased or normal PC in brain samples of people with LOAD.^39^ Similar to findings with PC, phosphatidylethanolamine (PE), phosphatidylinositol (PI), and phosphatidylserine (PS), along with phosphatidic acid (PA), have all been reported to be decreased in post-mortem brains from people with LOAD.^3, 21^ As a whole, the results suggest that most glycerophospholipids are decreased in tissue samples from people diagnosed with LOAD.

Lysoglycerophospholipids have only one FA side chain. They have been less studied than glycerophospholipids, which have 2 FA side chains. Lysophosphatidylcholine (LPC) was observed to be abnormally high in a postmortem study of LOAD, and LPC correlated positively with Aβ content of brain.^4^ In CSF, LPC, individually, and the ratio of LPC/PC were low in people with LOAD.^40^ In plasma, one study observed that both LPC and PC, individually, as well as the ratio of LPC/PC were low in people who concerted to LOAD.^41^

Phosphatidyl glycerol (PG), which has a glycerol backbone and a charged head group, but no FA side chains, has been observed to be elevated in post-mortem studies of MCI and LOAD.^4^ Levels of PG were strongly associated with numbers of plaques and tangles in brain.

Diacylglycerol (DAG), a glycerophospholipid with two FA side chains and no charged head group, was reported to be significantly elevated in plasma and post-mortem brains of people with MCI or early LOAD.^23, 32, 42^

### Sphingolipids in LOAD

Sphingolipids, like glycerophospholipids, are a class of compounds with varying FA side chains and polar head groups. Examples include sphingomyelin, found in many cell membranes; ceramides, which are part of cell membranes and also act as second messengers; and gangliosides, a large family of glycosphingolipids highly represented in lipid rafts within plasma membranes of cells in the brain, where they modulate the activity of ion channels.^43^ Once again illustrating the many roles of lipids, gangliosides also serve as recognition and communication molecules between cells.^44^

Studies on postmortem tissue show altered profiles of sphingolipids in LOAD.^23^ However, the studies do not consistently agree on which sphingolipids are found at higher and lower levels. ^3, 43^ This may be because sphingolipid levels change both with age and through the course of illness.^39^ The entire class is highly interactive with other factors observed to be abnormal in LOAD, including other lipids.^45^ In addition, they are reactive to stress and immune functions. ^27^ Studies of ceramides, in particular, suggest that levels may be increased early in illness and decreased at later times.^4, 27^

### Fatty Acids in LOAD

FA in brain and periphery exist either as side chains, conjugated to other lipids, or as free FA. The latter are stored in lipid droplets or are used as signaling molecules within the cell. In most studies of people with LOAD, no analyses to determine the relative amount or proportion of different FA types were performed or reported. When studied, FA, as a group, appeared to be low in post-mortem samples of brain from people with LOAD,^4, 46^ with the possible exception of docosahexaenoic acid (DHA).^46^ In lipid rafts isolated from post-mortem brain, polyunsaturated FA, including DHA, were low in samples from people with LOAD.^47^ In a post-mortem study of FA attached to phosphatidylcholines, all classes of FA were observed to be abnormally low, with unsaturated FA being particularly low.^48^ Aside from the general observation that FA are low in post-mortem brains in LOAD, there is no current agreement on which group or species of the numerous FA found in brain are most altered in LOAD.^4, 23^

## METHODS

### Overall design and hypotheses

The results presented in this paper are from studies of cell lines from patients with LOAD compared to similarly grown lines from healthy individuals. The lines were all reprogrammed iPSCs differentiated to astrocytes. Not all lipid abnormalities may be generally shared across different cell types. We analyzed astrocytes, as a first choice, because they have all relevant cell membranes and organelles, use lipids as signaling molecules, and have a role in inflammation, all of which may be altered in LOAD. They are involved in synapse integrity, protection against excitatory neurotoxicity, and maintenance of the blood-brain barrier.^49^ Notably, astrocytes provide metabolites for energy metabolism and cholesterols for membrane maintenance to neurons. ^21^ These properties make them highly relevant to risk for LOAD.

For a general estimate of cell lipids, whole cell lipid content was analyzed. In addition, recognizing that different organelles have somewhat different lipid constituents,^20^ mitochondria were isolated from the astrocytes and their membrane contents were analyzed. Mitochondria, as a subcellular component, were chosen for these first studies because mitochondrial membrane composition can profoundly affect bioenergetic flux,^50^ and bioenergetic abnormalities have been observed as prominent features in brains and cells from individuals with LOAD.^22, 51^

Based on results from previous studies, summarized above, it would appear that there are alterations in cholesterols, and may be alterations in glycerophospholipids and their components, as well as in sphingolipids and FA in association with LOAD. Among cholesterols, it appears that levels of CE are low, *in vivo,* and that such levels are predictive of illness. Genetic associations, and the specific association of APOE variants with LOAD, also suggest a role for cholesterol species, including CE, as being mechanistic determinants of risk for LOAD. On those bases, exploring whether CE levels are inherently abnormal may fairly represent the primary hypothesis. Among glycerophospholipids, the total of such lipids and the ratio of LPC/PC appeared to be abnormal in LOAD. Studies in reprogrammed cells in culture can determine if that abnormality is inherent. Regarding sphingolipids, the current evidence is less clear, but species within the class may be up or down in association with LOAD. Their levels are worth exploring in derived cell lines, but a prior hypothesis cannot be posed. Finally, the studies reviewed above suggest that there may be an association of low FA and LOAD.

### Subjects

All experiments and procedures were performed in accordance with relevant institutional and federal guidelines and regulations and were approved by the Mass General Brigham Institutional Review Board with written informed consent provided by all subjects. Subjects (Table 1) were recruited from the McLean Hospital Memory Diagnostic Clinic and diagnosed by a geriatric psychiatrist using the Diagnostic and Statistical Manual of Mental Disorders (DSM-5) criteria,^52^ using procedures including completion of the Montreal Cognitive Assessment (MOCA) test.^53^ When clinically indicated, additional neuropsychological testing was conducted to clarify diagnosis. All subjects were assessed for their capacity to provide consent. In the case of incapacity, an authorized guardian/healthcare proxy provided consent, and the subject provided assent.

**Table 1.** Subject Demographics.

|  | **HC** | **LOAD** |
| --- | --- | --- |
| **Number of Subjects** | 11 | 11 |
| **Age (yrs)** | 52.1 +/- 21.8 | 72.9 +/- 9.5 |
| **Sex** | Male: 8, Female: 3 | Male: 6, Female: 5 |
| **APOE** | E2/2: 1<br>E2/3: 1<br>E3/3: 7<br>E3/4: 2 | E3/3: 1<br>E3/4: 8<br>E4/4: 2 |
| **MOCA Score** | - | 13.4 +/- 7.0 |

HC: Healthy Control; LOAD: Late-Onset Alzheimer's Disease; APOE: Apolipoprotein ε; MOCA: Montreal Cognitive Assessment.

### Tissue culture

Establishment of the iPSC lines and culture protocols were as described in previous publications.^51, 54, 55^ Fibroblasts or peripheral blood mononuclear cells (PBMCs) donated by subjects were reprogrammed to iPSCs by standard protocols. At passage numbers of 18 or higher, iPSCs were differentiated to neural progenitor cells (NPCs) by chemical neural induction methods. NPCs were propagated and characterized at passage 5 for the expression of NPC markers, including NESTIN, PAX6, and SOX2, detected by fluorescence immunocytochemistry (FICC). NPCs were further differentiated to astrocytes using a 30-day chemical differentiation protocol and characterized for the expression of astrocytic markers, including GFAP, GLAST, and S100ß.

### Mitochondria extraction

Nine million astrocytes were used for mitochondrial extraction utilizing a Mitochondria Isolation Kit for Cultured Cells (Thermo Scientific, Cat No. 89874) following the manufacturer’s instruction with slight modifications according to Kappler^56^, Preble^57^, Williamson^58^ and Kurokin.^59^ To minimize thermal effects on mitochondrial integrity, all reagents, instruments, and intermediates of mitochondrial extracts were kept on ice during the entire process, and the temperature of the centrifuge was set at 4 °C. Cells were lysed with Reagent A from the kit, with a brief vortex followed by incubation on ice for 2 mins and subsequent homogenization using a KIMBLE Dounce Tissue Grinder (DWK life sciences, Wertheim, Germany, Cat No. 885300-0002). Reagent C from the kit was added to the homogenized cell lysates and the mixture was centrifuged to remove cellular debris. Astrocyte lysates were centrifuged in two steps, at 1,000 g for 5 mins and 1,300 g for 10 mins. Supernatants containing the cytosolic and mitochondrial fractions were then partitioned for further processing: About 1/3 for Western blots and 2/3 for lipidomic analysis. Separation into cytosolic fractions, as supernatants, and mitochondria, as pellets, was achieved with 15 mins centrifugation at 3,000 g. After a washing step with Reagent C, mitochondrial pellets for Western blots were solubilized in 2 % 3-[(3-Cholamidopropyl)dimethyl-ammonio]-1-propane sulfonate (CHAPS, Roche, Basel, Switzerland, Cat No. 10810118001) dissolved in 1 X Tris-Buffered Saline (TBS, Boston BioProducts Inc, MA, USA, Cat No. BM-301X) in distilled water, vortexed for 1 min, and centrifuged at 15,000 rpm for 2 mins to obtain supernatants as a final mitochondrial fraction. Mitochondrial pellets used for lipidomic analysis were washed with Reagent C and resuspended in 1 X DPBS without magnesium and calcium and stored at-80 °C until further use.

### Western blots

Proteins were quantified by the Pierce bicinchoninic acid (BCA) assay (Thermo Scientific, Cat No. 23225), with approximately one million astrocytes lysed in RIPA Lysis and Extraction buffer (Thermo Scientific, Cat No. 89900) used as positive controls.

Twenty-five micrograms of whole cell lysates or mitochondrial extracts were mixed with Western-Ready Protein Sample Loading Buffer (BioLegend, Cat No. 426311) containing 5 % beta mercaptoethanol (ß-ME, Sigma-Aldrich, Cat No. M3148) and heated at 60 °C for 10 mins. Denatured samples were run on 12 % sodium-dodecyl-sulfate polyacrylamide gel electrophoresis (SDS-PAGE) gels at 100 V for about 1.5 hours. Gels were then blotted on polyvinylidene difluoride (PVDF) membranes (Bio-Rad, CA, USA, Cat No. 1620177) at 100 V for 2 hours, followed by membrane blocking with 5 % skim milk (Bio-Rad, Cat No. 1706404) in 1 X TBS in distilled water with 0.1 % TWEEN-20 (Sigma-Aldrich, Cat No. P1379) at RT for one hour. Several subcellular organelle markers^56^ were used to determine the purity of the mitochondrial extracts (Supplementary Table S1, Supplementary Fig. S1). Hybridized primary antibodies were captured by horse radish peroxidase (HRP) conjugated secondary antibodies (Vector Laboratories; Cat No. PI-2000-1, RRID: AB_2336177; and Cat No. PI-1000-1, RRID: AB_2336198) at RT for one hour followed by treatment with West-Q Femto Clean electrogenerated chemiluminescence (ECL) solution (GenDEPOT, TX, USA, Cat No. W3680-020) and signals were measured using a ChemiDoc MP imaging system (Bio-Rad, RRID: SCR_019037).

### Lipidomics analysis

Untargeted shotgun lipidomic analyses were performed by Lipotype (Dresden, Germany). Whole cells and mitochondrial extracts were subjected to lipid extraction using a chloroform/methanol method.^60^ Spiked with internal lipid standards, lipid extracts were analyzed with a Q-Exactive mass spectrometer equipped with a TriVersa NanoMate ion source (Advion Bioscience, NY, USA) both in positive and negative ion modes.^61^ Individual species of 33 lipid classes were detected (Supplementary Table S2). A detailed description of lipidomic profiling by Lipotype is presented in Liebisch^62^ and Summa.^63^

### Data Processing

Lipid species were identified based on molecular mass using LipotypeXplorer (Lipotype)^64^ Identified lipid species were primarily quantified by peak areas, which were filtered by signal-to-noise and signal-to-background ratios to sort out false signals: Lipid species whose ratios were lower than 5 were filtered out. Filtered lipid species in peak areas were then converted to lipid amounts in pmol units using calibration curves established by internal lipid standards. Prior to downstream analysis, data filtering was applied using an occupational threshold of 0.7;^65, 66^ that is, lipid species that were measured in more than 70 % of either control subjects or LOAD patients were kept in the analysis. Also consistent with standard analyses, missing values; that is, lipid species undetected in individual samples at the thresholds obtainable, were imputed by half the minimum value of detection for each lipid species. Lipid amounts were calculated as mol% of total lipids, and lipid species data were summed within lipid classes. The results from whole cells and mitochondria were processed independently. Data processing was performed in R (version 4.4.2; RRID: SCR_001905).

### Statistics

Unless other stated, two-tailed Students’ t-tests, uncorrected for multiple measures, were used to compare measurements of lipids between samples from individuals with LOAD and healthy control subjects. Analyses were performed in Microsoft Excel for Mac, version 16.96.1, and PRISM 9 for macOS, version 14.1. (GraphPad). Differences were considered statistically significant when *p*-values were less than 0.05, while *p*-values between 0.05 and 0.1 were considered as a trend towards significance. Lower CE, consistently associated with LOAD in CSF and blood studies, was treated as a prior hypothesis.

## RESULTS

Results are presented for lipid contents in whole cell preparations or mitochondria. Differences are noted as percent changes of lipid levels in cell lines derived from individuals with LOAD compared to healthy control subjects. For comparison to results from post-mortem, CSF, and plasma studies, data were analyzed by classes of lipids.

### Lipid Levels in Whole Cells

After data filtration and imputation (see Materials and Methods), a total of 1,047 lipid species were detected in astrocytes, among which, 684 (65.3%) were lower and 363 (34.7%) were higher in cells from individuals with LOAD. No prior hypothesis was proposed regarding results at the species level. Rather, as detailed above, hypotheses were based on prior studies that examined lipids by class. A summary of the results by lipid classes is given in Table 2, and details of results are described below.

**Table 2.** Lipid classes detected in whole cell astrocyte preparations.

| **Lipid Class** | **HC** | **SEM** | **LOAD** | **SEM** | **% LOAD** | **% SEM** | ***p* value** |
| --- | --- | --- | --- | --- | --- | --- | --- |
| **CE** | 1.40 | 0.26 | 0.66 | 0.22 | 47.02 | 15.59 | 0.021 |
| **Cer** | 0.51 | 0.04 | 0.54 | 0.05 | 105.94 | 10.78 | 0.660 |
| **Chol** | 26.53 | 1.38 | 27.32 | 1.33 | 102.97 | 5.02 | 0.686 |
| **DAG** | 3.49 | 0.24 | 2.99 | 0.26 | 85.73 | 7.45 | 0.174 |
| **DiHexCer** | 0.09 | 0.01 | 0.13 | 0.02 | 149.31 | 25.39 | 0.108 |
| **GM3** | 0.50 | 0.09 | 0.55 | 0.10 | 109.69 | 18.88 | 0.714 |
| **HexCer** | 0.37 | 0.03 | 0.38 | 0.03 | 104.49 | 8.32 | 0.708 |
| **LPA** | 0.06 | 0.02 | 0.11 | 0.02 | 182.91 | 37.18 | 0.098 |
| **LPC** | 0.21 | 0.04 | 0.30 | 0.10 | 141.52 | 47.15 | 0.422 |
| **LPC O-** | 0.06 | 0.01 | 0.05 | 0.01 | 86.82 | 20.66 | 0.650 |
| **LPE** | 0.25 | 0.04 | 0.26 | 0.05 | 103.87 | 20.23 | 0.886 |
| **LPE O-** | 0.26 | 0.07 | 0.19 | 0.04 | 73.34 | 16.69 | 0.386 |
| **LPG** | 0.02 | 0.00 | 0.01 | 0.00 | 87.18 | 8.16 | 0.279 |
| **LPI** | 0.05 | 0.02 | 0.05 | 0.01 | 93.33 | 23.92 | 0.882 |
| **LPS** | 0.08 | 0.02 | 0.07 | 0.01 | 91.18 | 15.50 | 0.779 |
| **PA** | 0.42 | 0.07 | 1.48 | 0.44 | 352.71 | 106.17 | 0.029 |
| **PC** | 31.63 | 1.09 | 30.33 | 1.21 | 95.89 | 3.81 | 0.434 |
| **PC O-** | 4.64 | 0.46 | 4.45 | 0.36 | 95.88 | 7.68 | 0.745 |
| **PE** | 7.54 | 0.31 | 7.87 | 0.25 | 104.45 | 3.35 | 0.413 |
| **PE O-** | 6.53 | 0.40 | 5.74 | 0.25 | 87.93 | 3.81 | 0.111 |
| **PG** | 1.17 | 0.10 | 1.00 | 0.08 | 86.05 | 6.64 | 0.204 |
| **PI** | 4.29 | 0.45 | 4.99 | 0.16 | 116.55 | 3.79 | 0.157 |
| **PS** | 7.48 | 0.44 | 7.83 | 0.38 | 104.64 | 5.12 | 0.558 |
| **SM** | 2.27 | 0.09 | 2.53 | 0.16 | 111.76 | 7.11 | 0.167 |
| **TAG** | 0.16 | 0.03 | 0.15 | 0.05 | 91.66 | 33.65 | 0.833 |

Numbers depict average mol% (HC and LOAD), and percent levels in LOAD over HC-associated samples. *p* values are calculated by two-tailed t-tests, except for CE, for which a one-tailed t-test was used. HC: Healthy controls; LOAD: Late-onset Alzheimer's disease; SEM: Standard error of the mean.

*Levels of Cholesterol and Cholesterol Esters:* Neutral cholesterols were not different in cells derived from individuals with LOAD (103%, SEM=5%). Levels of CE, however, were quite different between groups. Five CE passed the filtration criteria and as a class were 53% lower in the LOAD group than in cells from the healthy comparison group.

Reduced CE in cells from individuals with LOAD is a prior hypothesis. Therefore, a one tailed t-test, uncorrected for multiple measures, is appropriate, giving a p=0.021 (with SEM=15.6%), for the deficit in CE observed in these cells. While significantly lower CE was observed in CSF and blood of people with LOAD, those studies did not report data on which species within the lipid class CE were low. Different species of CE have different FA side chains, and these were measurable by the methods applied in this study. Notably, all five detectable CE species were substantially decreased in cells from individuals with LOAD. Decreases for individual species of CE ranged from 49-62%, with p values ranging from 0.015 to 0.028. (Table 3).

**Table 3.** CE in whole cell astrocyte preparations.

| **CE** | **HC** | **SEM** | **LOAD** | **SEM** | **% LOAD** | **% SEM** | ***p* value** |
| --- | --- | --- | --- | --- | --- | --- | --- |
| **16:0;0** | 0.28 | 0.05 | 0.14 | 0.05 | 51.33 | 16.61 | 0.028 |
| **16:1;0** | 0.15 | 0.03 | 0.07 | 0.03 | 46.62 | 17.04 | 0.024 |
| **18:0;0** | 0.14 | 0.03 | 0.05 | 0.02 | 38.12 | 12.13 | 0.015 |
| **18:1;0** | 0.72 | 0.14 | 0.34 | 0.12 | 46.81 | 16.12 | 0.026 |
| **18:2;0** | 0.12 | 0.02 | 0.06 | 0.02 | 49.17 | 16.87 | 0.026 |

Numbers depict average values in mol% (HC and LOAD), and percent lipid levels in LOAD over HC-associated samples. *p* values are calculated by one-tailed t-test. HC: Healthy controls; LOAD: Late-onset Alzheimer's disease; SEM: Standard error of the mean.

APOE variants may differentially transport CE. Therefore, evidence of a possible association of APOE4 genotype and CE level was explored in the sample populations. Of the 11 healthy comparison individuals, 2 were heterozygote carriers of APOE4 and one of the patients with LOAD carried an E3/3 haplotype (Table 1). When the groups were clustered by APOE3 or E4 genotype, CE, as a lipid class, was 27% (SEM=20.2%, p=0.194) lower in the APOE4 carrier group, suggesting that APOE4 may contribute to low CE levels in the cells from people with LOAD. However, as most of the individuals with APOE4 genotypes were individuals with LOAD, this difference may reflect other factors leading to risk for LOAD. Therefore, results were examined separately within the LOAD and healthy comparison groups. The two samples from people in the healthy comparison group who carried an E4 haplotype had CE values of 2.19 and 1.34 mol%, averaging 1.76 mol%, which was similar to the whole healthy control group average of 1.4 mol%. The cells from the individual with LOAD genotyped as APOE3/3 had CE levels of 0.21 mol%, even lower than the average for the whole group with LOAD, at 0.66 mol%, or the entire group with an E4 haplotype, at 0.88 mol%. Taken together, the data did not suggest an association of APOE4 genotype with lower CE. Rather, it suggested a direct association of low CE with LOAD.

*Glycerophospholipids and their derivatives: Glycerophospholipids* in astrocytes derived from iPSC did not follow the patterns observed in post-mortem studies. Contrary to the decreases observed in post-mortem analyses of the brain in LOAD, PA appeared to be substantially elevated, by 253% (SEM=106.2%), in the LOAD group versus the healthy comparison group, with p=0.029 for the difference. LPA was also elevated, by 83% (SEM=37.2%), trending towards significance at p=0.098. PC was detected with and without esterified side chains. Both classes were modestly, but non-significantly, decreased. LPC without an esterified side chain was increased 42%, and LPC with an esterified side chain was decreased 13%, with neither difference reaching statistical significance. The ratio of LPC/PC was 35% higher in the LOAD samples than the controls, but that did not reach statistical significance (SEM=41.7%, p=0.45). DAG appeared to be decreased in the LOAD group (down 14%, SEM=7.5%) versus the healthy comparison group, and the difference was not significant (p=0.174). This decrease in DAG is opposite to increases observed in studies of post-mortem brain and plasma in people with LOAD. Triacylglycerol (TAG) was also slightly reduced, by 8%, but that deficit was not statistically significant (p=0.833).

*Sphingolipids:* Neither ceramides, which show changes over course of illness in post-mortem studies of LOAD, nor sphingomyelin appeared to be much different between cells derived from individuals with LOAD and cells from healthy controls. There were no differences approaching nominal significance nor even notable trends towards group differences of sphingolipids.

*Fatty acids*: Individual FA were more likely to be decreased in cells derived from individuals with LOAD (see Figure 1a). For the entire group, the reduction averaged 6% (SEM=7.3%, p=0.44). Of 56 species detected, 14 (25%) were saturated FA, and these were decreased an average of 9% (SEM=9.4%, p=0.273) in cells in the LOAD group.

**Figure 1.**
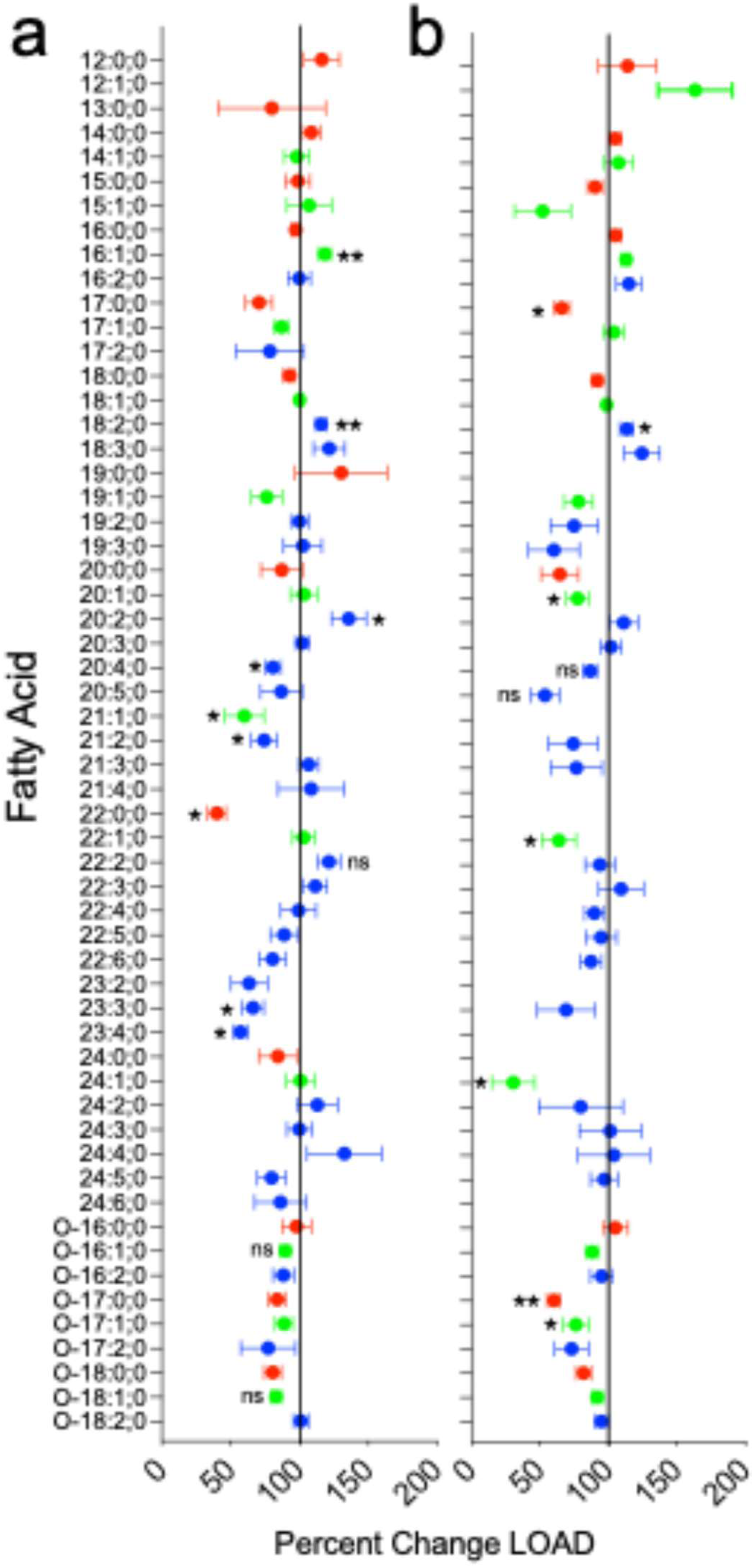
Fatty Acids. FA in (a) whole cell astrocyte preparations or (b) isolated mitochondria preparations plotted as percent lipid levels in LOAD over healthy control associated samples using mol% values. Red: Saturated FA; green: Monounsaturated FA; blue: Polyunsaturated FA. Data were analyzed by two-tailed t-tests. FA levels are labeled where the difference between values in LOAD and HC associated samples reached a trend towards significance p < 0.1 (ns), or significance, uncorrected for multiple measures, *p* < 0.05 (*), or *p* < 0.02 (**). Unlabeled differences did not reach a p value less than 0.1.

The largest decrease was seen for docosanoic acid (22:0), which was reduced 60% (SEM=6.7%, p=0.046). There were 13 monounsaturated FAs (23% of all FA species detected), and as a group, they were decreased by 6%, (SEM=3.9%, p=0.39). The greatest decrease observed was in heneicosenoic acid (21:1), which was reduced 40% (SEM=14.4%, p=0.049). There were 29 polyunsaturated FAs (52% of all species detected), and as a group they were decreased by 4% (SEM=3.8%, p=0.352). Among the polyunsaturated FA species, several were substantially reduced, including two members of the docosatetraenoic acid group, 23:4, down 43% (SEM=4.7%, p=0.020), and 23:3, down 34% (SEM=8%, p=0.035). Arachidonic acid (20:4), which plays a role in inflammation, ^67^ was down 19% (SEM=5.8%, p=0.027) in the LOAD group. More details on trends in individual species, many of which are decreased in the LOAD group, are shown in Figure 1a.

### Lipid Levels in Mitochondria

In mitochondria isolated from astrocytes, a total of 448 lipid species were detected after data filtration and imputation, of which 341 were lower (76.1%) and 107 (23.9%) were higher in samples from individuals with LOAD. As with whole cell results, the lipid species were grouped by class (Table 4).

**Table 4.** Lipid classes detected in astrocyte mitochondria preparations.

| **Lipid Class** | **HC** | **SEM** | **LOAD** | **SEM** | **% LOAD** | **% SEM** | ***p* value** |
| --- | --- | --- | --- | --- | --- | --- | --- |
| **Cer** | 0.26 | 0.04 | 0.20 | 0.02 | 78.62 | 5.81 | 0.204 |
| **Chol** | 31.17 | 1.41 | 32.07 | 1.29 | 102.89 | 4.13 | 0.642 |
| **DAG** | 2.74 | .38 | 2.83 | 0.18 | 103.00 | 6.57 | 0.848 |
| **GM3** | 0.88 | .15 | 1.03 | 0.15 | 116.99 | 17.10 | 0.489 |
| **HexCer** | 0.35 | 0.04 | 0.32 | 0.03 | 90.68 | 8.03 | 0.506 |
| **LPC** | 0.12 | 0.04 | 0.05 | 0.01 | 46.60 | 5.84 | 0.096 |
| **LPE** | 0.14 | 0.04 | 0.08 | 0.01 | 58.41 | 5.02 | 0.141 |
| **LPE O-** | 0.07 | 0.02 | 0.03 | 0.00 | 43.09 | 3.54 | 0.088 |
| **PA** | 0.09 | 0.01 | 0.11 | 0.03 | 121.69 | 33.25 | 0.535 |
| **PC** | 31.81 | 1.02 | 31.85 | 0.65 | 100.15 | 2.05 | 0.969 |
| **PC O-** | 4.83 | 0.47 | 4.87 | 0.29 | 100.81 | 6.05 | 0.945 |
| **PE** | 8.03 | 0.50 | 8.42 | 0.28 | 104.90 | 3.47 | 0.499 |
| **PE O-** | 6.99 | 0.42 | 6.04 | 0.45 | 86.43 | 6.44 | 0.141 |
| **PG** | 1.09 | 0.06 | 0.84 | 0.05 | 76.98 | 4.66 | 0.006 |
| **PI** | 2.37 | 0.12 | 2.30 | 0.11 | 97.12 | 4.62 | 0.675 |
| **PS** | 6.01 | 0.25 | 5.94 | 0.32 | 98.79 | 5.29 | 0.858 |
| **SM** | 3.06 | 0.17 | 3.01 | 0.13 | 98.55 | 4.40 | 0.840 |

Numbers depict average mol% values (HC and LOAD), and percent levels in LOAD over HC-associated samples. *p* values are calculated by two-tailed t-test. HC: Healthy controls; LOAD: Late-onset Alzheimer's disease; SEM: Standard error of the mean.

Mitochondrial membranes have a more restricted composition than other cell membranes and not all the lipids and differences observed in mitochondria are the same as those observed in the whole cell preparations.

*Levels of Cholesterol:* As observed in results from analyzing whole cell astrocytes, cholesterol was not different in mitochondria isolated from cells derived from people with LOAD and control individuals (13% increase, SEM=4.1%, p=0.642). Most CE are stored in separate organelles, and the low amounts of CE in mitochondria were below the level of detection in the Lipotype assays.

*Glycerophospholipids and their derivatives:* The only glycerophospholipid that had substantially different levels in mitochondria from the cells derived from people with LOAD was phosphatidylglycerol (PG), which was decreased by 23% (SEM=4.7%, p=0.006). This is opposite to observations of PG in cortex samples in post-mortem studies of LOAD. Of course, regional brain samples contain many other cellular elements and their lipids, not just mitochondria.

Among other glycerophospholipids, most showed no substantial difference between groups. However, lysoglycerophospholipids were generally lower in mitochondria from individuals with LOAD. All lysoglycerophospholipid types were lower, by 50-60%, with LPC reduced 53% (SEM=5.8%, p=0.096). The ratio LPC/PC was down by 54% (SEM=6.13%, p=0.095). Unlike the results in the whole cell preparation, DAG was not decreased in mitochondria from the LOAD group.

*Sphingolipids:* Ceramides in mitochondria from the LOAD group trended low, being 23% down for the entire class, but these differences did not reach statistical significance (p=0.204). However, adding to the suggestion of a deficit, there were 6 species of ceramide detected, each with a different fatty acid side chain, and every species was lower in the LOAD group. Although none of these deficits reached statistical significance, the finding that they were all reduced suggests the possibility of a deficit in ceramides in mitochondria in LOAD.

*Fatty acids*: Fewer species of FA were detected in mitochondria than in whole cells, but the overall patterns of differences in FA species between groups in the mitochondria preparation were quite similar to the differences seen in the whole cell preparations, see Figure 1b. Thus, as in the whole cells, FA, on average, were decreased, down 10% (SEM=3.4%, p=0.278), in the LOAD group. Among 47 FA species detected, there were 10 (21%) species of saturated FA. Most were reduced in the LOAD group, and the whole group was down 12% (SEM=8.1%, p=0.30). The highest significant difference was reached for heptadecanoic acid (17:0), down 34% (SEM=6.0%, p=0.032) and its ester, down 40% (SEM=5.0%, p=0.088). There were 13 (28%) monounsaturated species detected, which were decreased 12% on average for the whole group (SEM=9.6%, p=0.265). Individual monounsaturated species with decreases that were statistically significant or nearly significant included eicosenoic acid (20:1), down 23% (SEM=8.2%, p=0.047), heptadecenoic acid ester (0-17:1), down 24% (SEM=9.4%, p=0.055), docosenoic acid (22:1), down 36% (SEM=13%, p=0.063), and nervonic acid, so named because it was first discovered in mammalian nervous tissue and is essential for brain health^68^ (24:1), down 69% (SEM=16%, p=0.079). One monounsaturated species, palmitoic acid (16:1) was increased 13% (SEM=3.6%, p=0.017), and dodecenoic acid (12:1) was increased 63% (SEM=27%, p=0.159) in the LOAD group.

There were 24 (51% of all species) that were polyunsaturated FA. On average, they were decreased by 9% (SEM=9.4%, p=0.46). Most individual species were down, but none of these decreases reached statistical significance. The greatest deficit was in eicosapentaenoic acid (20:5), an omega-3 fatty acid involved in inflammation.^69^ It was down 46% (SEM=10.1%, p=0.091).

## DISCUSSION

### Context of the studies

Findings on post-mortem tissue, along with results from CSF and plasma studies, reflect changes associated with progression and treatment of LOAD, as well as aging and any associated illness. Post-mortem studies, in particular, often represent disease endpoints. Results from studies of reprogrammed cells, lacking markers of aging or disease, as reported here, are complementary to post-mortem, CSF, and plasma studies in that they can detect inherent factors. Taken together, clinical studies and reprogrammed cell culture studies may both identify and separate mechanisms underlying neuropathological risk factors and progression of illness.

To our knowledge, no previous studies have determined whole lipid contents in cells obtained from people with LOAD reprogrammed to iPSCs and then differentiated to become brain cells. We believe that these are the first results that provide those data for comparison to other approaches determining lipid levels associated with LOAD.

There are a few studies using isogenic APOE4 iPSC lines that study the relationship of APOE variants to some aspects of lipid metabolism. Generally, these are not studies of lines from people with LOAD. Nevertheless, they are worth noting. One past study observed that iPSC-differentiated APOE4 isogenic astrocytes had a 20% increased *de novo* synthesis of total cholesterol, but not cholesterol esters, as well as increased cholesterol accumulated in lysosomes when compared to the same APOE3 haplotype cell line. ^70^ Other studies on isogenic APOE4 iPSC lines confirm that APOE4 is associated with altered metabolism of cholesterols and FA, as well as increased formation of lipid droplets.^71, 72^ In a study of iPSC from individuals with LOAD differentiated to cortical neurons, dysregulation of genes involved in lipid metabolism were observed, but lipid levels were not measured.^73^ Further work combining characterization of lipid levels, as performed here, and of underlying mechanisms, including the role of APOE4, should lead to the identification of specific aspects of lipid composition and lipid metabolism that increase the risk for LOAD.

### Review of the findings

The findings of substantial deficits of CE in astrocytes from people with LOAD are consistent with similar abnormalities reported *in vivo* in blood and CSF studies (see ^21, 24, 30, 31, 33^). By comparison, levels of cholesterol, per se, were not substantially or statistically significantly different in cells from people with LOAD and healthy people.

Similarly, in clinical studies of LOAD, no substantial abnormalities of cholesterol levels were observed.^24^ It is possible that the modest decreases in cholesterol observed here, in cells from people with LOAD, are related to the decreases in cholesterol synthesis noted in clinical studies of LOAD.^15, 16^

Contrary to findings from post-mortem studies, as reviewed above, substantial decreases of PC or most other glycerophospholipids were not seen, with the exception that PG was low both in whole cell astrocyte preparations and mitochondria. PA, reported along with other glycerophospholipids to be decreased in post-mortem studies of LOAD,^3, 21^ was increased in whole cell preparations, though not in mitochondria, in the samples from people with LOAD studied here. Evidence that the ratio of LPC/PC was abnormal, and even opposite in whole cells versus mitochondria, was notable but inconclusive.

It may not be surprising that major lipids, specifically, cholesterols and glycerophospholipids that are present at high concentrations and used structurally in cell membranes, were similar in the two groups. Large changes in cholesterol and some glycerophospholipids, such as PC, would greatly compromise many, if not most cell functions, and might be expected to be associated with substantial developmental disorders or the early onset of neurodegeneration. Therefore, even if there are genetically determined abnormalities in the metabolism of these classes of lipids, there may also be compensatory mechanisms that maintain the lipids at roughly normal levels, at least in early life. Findings of large changes in glycerophospholipids in post-mortem studies could be associated with cell disintegration, as occurs in established disease.

While major lipids are largely structural, CE is a signaling and storage molecule, present at levels an order of magnitude lower than those of neutral cholesterols, and the effects of abnormalities in CE levels may be better tolerated, causing less dysfunction in cell processes. Similarly, abnormal levels and metabolism of a few glycerophospholipids, such as PG or PA, as seen here, or in LPC/PC, might be better tolerated than deficits in all glycerophospholipids as a class, as observed post-mortem in LOAD.^24^

Nonetheless, CE is necessary, as a precursor, for maintaining cholesterols in cell membranes, and altered cholesterol precursor levels found in CSF and plasma of patients has suggested that cholesterol *de novo* synthesis is reduced in LOAD. ^16^ CE levels decrease naturally with age,^74^ and inherently low CE as seen in cells from patients with LOAD might be a risk factor for inadequate membrane maintenance over the course of a long life. In fact, CEs may play a key role in a cascade of effects underlying illness progression.^23^ Specifically, *in vitro* studies observed that higher CE is associated with decreased production of toxic Aβ oligomers and lower CE with increased synaptic damage from Aβ fragments.^75–77^ And decreased production of cholesterols, typically dependent on the levels of CEs, increases hyperphosphorylation of Tau protein. ^78^ By these mechanisms, the lower CE observed here in the cells from people with LOAD may directly contribute to the production and effects of toxic abnormal proteins seen in many cases of LOAD.

FA of all subtypes were consistently, though modestly, down in the LOAD group studied here, in both whole cells and mitochondria. This is consistent with results from clinical studies of FA in association with LOAD.^4, 46, 48^ Some species were reduced more than others. Only a few were elevated. Overall, the findings suggest imbalances in FA content associated with LOAD. As with CE, these reductions and imbalances could compromise many cell functions but might well be tolerated during development and early life, only leading to an increased risk of illness over time. Abnormal concentrations of FA in glycerophospholipids, would affect cell membrane fluidity^21^ and thereby alter the processing of proteins, including amyloid precursor protein (APP). Membrane fluidity abnormalities have been proposed elsewhere as a mechanism underlying the production of toxic Aβ oligomers.^79^ In addition, as signaling molecules, FA are involved in cell maintenance functions and in inflammation. FA reduction, especially reduction in key FA species, might impair the response of cells to stress and aging.^80^ And FA can serve as stores of substrates for energy production through β-oxidation, another function that is compromised in those at risk for LOAD.

What may explain inherent differences in cell lipid contents? There may be abnormalities of anabolism and/or catabolism. The ER has been noted as the site where proteins are processed and where APP is misprocessed. And the ER is where lipid droplets, containing CE and FA, are produced.^8^ Abnormal internal membrane proportions,^81^ particularly in the ER,^82^ have been reported in LOAD. Lower CE content might be a consequence of abnormal ER function. In addition, lipids are key components of phagosomes, endosomes, and lysosomes, thus contributing to the processing, digestion, and elimination of proteins and lipids by endocytosis and autophagy.^8^ Reciprocal interactions have been observed between lipids and Aβ peptides in LOAD (and EOAD), and these interactions involve the formation and activity of cellular organelles.^83–85^ This would include mitochondria, as noted. Inherent abnormalities of bioenergetics have been documented in LOAD.^22, 51^ Altered endocytosis and autophagy have also been observed as inherent properties of cell lines from individuals with LOAD.^86, 87^ Overall, the evidence suggests that abnormalities of both anabolism and catabolism of lipids may be inherent factors underlying risk for LOAD.

In his original paper on a case of dementia, Alois Alzheimer noted lipid accumulations.^88^ This observation has been consistently confirmed in subsequent research on both early and late onset AD, with a specific increase in lipid droplets and some of their contents.^8, 27^ By comparison, in the reprogramed cell lines studied here, decreases in CE and FA, both major contents of lipid droplets, were observed. PA, a precursor in FA synthesis, was observed to be increased. DAG, and possibly TAG, both downstream of PA, were observed to be decreased. This suggests a metabolic abnormality in FA metabolism at the step from PA to DAG and TAG. The relationship between these inherent lipid deficits and the lipid deficits and accumulations observed post-mortem, requires further study.

### Limitations of the current study and opportunities for future work

Future studies with a greater number and diversity of cell lines are needed to confirm, refute, or expand this work. Other cells, including neurons, may not show the same abnormalities as astrocytes. Lines from more study participants are needed. This includes increasing the number of male and female participants, so that sex differences can be explored. The prevalence of LOAD is higher in women than men. This may be the result of hormonal influences, as well as sex-related differences in inflammatory activity.^89^ However, peripheral lipid metabolism differs in men and women, and this may contribute to risk for LOAD, as well.^89^ It is unknown if there are inherent differences in lipid metabolism in brain cells from men and women.

Although APOE variants were not clearly associated with the abnormalities observed in the lines from individuals with LOAD studied here, more studies are required to address the importance of APOE haplotypes on lipid content. Future studies will also be needed to determine whether the abnormalities seen here in astrocytes are also seen in other cell types.

Whole cell extracts contain lipids from a mix of membranes and organelles with different lipid compositions.^20^ The proportion of membranes in cells from people with LOAD may differ from those without LOAD. Specifically, the ratio of ER to other membranous elements may be higher in cells from individuals with LOAD. ^79^ This could affect the results observed in the whole cell preparation.

CE largely exist in lipid droplets, and it needs to be determined if lipid droplet amounts are increased or decreased in the LOAD cell populations analyzed here. Whether abnormal amounts or functions of the ER, specifically its production of lipid droplets, are related to abnormal levels of lipids observed here will also require further studies.

Mitochondria have an inner and an outer membrane, but the results reported here are for mitochondria as a whole. Overall, mitochondrial lipid composition in the astrocytes from patients with LOAD appears to be abnormal, but future studies will be needed to define abnormalities specific to various mitochondrial membranes.

Lipids in cells in culture come in part from the growth medium and in part from cellular metabolism of the nutrients the cells receive. Abnormal lipid levels observed in cells from people with LOAD may reflect differences in uptake of nutrients, synthesis of new lipids, or catabolism of lipids. All these mechanisms are ripe for future study.

Lipids are dynamic elements of cells. Profiling lipid levels doesn’t document lipid metabolism and interactions. However, observation of abnormalities can suggest leads for mechanistic studies. For example, the results reported here would suggest follow-up studies of the formation, interactions, and metabolism of CE in LOAD. Agents that alter CE metabolism and levels in blood are available.^90^ Drugs could be developed to modify CE levels and activities in brain. The same might be true for other abnormalities observed here. And lipids, themselves, can serve as restorative or therapeutic agents.^91^

Currently, tests to detect risk for AD, or for use in the diagnosis of AD, largely focus on determining Aβ and hpTau, or their derivatives, in plasma.^92^ These tests may be useful, especially for early diagnosis of those cases of LOAD associated with higher levels of plaques and tangles. However, the appearance of these compounds in blood may be secondary to other, more basic, predisposing mechanisms, and elevated levels of Aβ and hpTau in plasma or CSF may only be detectable after damage has already occurred in brain. In addition, many cases of LOAD are not associated with pronounced plaques and tangles, and many people who age without substantial cognitive deficits have high levels of plaques and tangles.^93^ Defining inherent cellular processes that contribute to early pathological processes and underlie the appearance of LOAD, rather than later stage elements of plaques and tangles, may lead to tests that can be applied long before brain damage has occurred. In addition, specific abnormalities observed in those tests, which may differ among individuals at risk, may detect subtypes of LOAD and suggest personalized approaches for interventions, including interventions targeting specific lipids and membranes.^22, 54, 94^ As an example from the results reported here, abnormalities of CE levels and synthesis, or processes underlying abnormalities of lipid droplet formation and contents, may be targets for beneficial modulation that could occur years or decades before the development of LOAD.

## Conclusion

Multiple approaches and sources, including genomic, epidemiologic, clinical, and post-mortem studies, observe a link between lipids and LOAD. Studies of cells from people with LOAD, as performed here, can identify which abnormalities, in which specific lipids, are inherently abnormal and may be risk-determining precursors for LOAD. The evidence from these new cell studies suggests that there are no prominent inherent abnormalities in levels of membrane structural lipids, including glycerophospholipids, sphingolipids, and cholesterols, associated with LOAD. Rather, the inherent abnormalities are in levels of storage and signaling lipids, most prominently CE, along with imbalances of FA. The findings have implications for understanding lipid-based risk factors for LOAD and for the design of studies to target and address those risk factors.

The findings reported here must be replicated and extended to include more cell lines and greater details on specific lipid abnormalities. They should be complemented by studies of mechanistic factors, such as specific metabolic reactions or aspects of lipid transport, that underlie inherent abnormal cell lipid composition associated with LOAD. And they should be complemented by further studies to evaluate how abnormal lipid composition affects cell functions. These results can be evaluated in the context of genomic and phenotypic characterizations of the same individuals who provide cell samples.

Despite being preliminary, the findings strongly suggest that further study of inherent lipid abnormalities associated with LOAD will be fruitful. Such studies should lead to greater understanding of mechanisms contributing to risk for LOAD and help explain the development of pathology observed in brain in LOAD. In time, the relationship of lipid abnormalities to other abnormalities predisposing to LOAD should become clear. New screening tests can follow. And most importantly, identifying key inherent factors associated with the risk for LOAD may lead to early interventions that improve the course of aging and reduce the risk of dementia later in life.

## Data Availability

All data produced in the present study are available upon reasonable request to the authors

## ACKNOWLEDEMENTS

Authors’ contributions: BMC conceptualized the study. BMC and KCS designed the study. EK performed the experiments. EK and KCS analyzed results, with input from BMC. KRL and IL contributed to analysis and interpretation of results. BMC wrote the paper, which was edited by BMC and KCS. The manuscript was read, commented on, and approved by all the authors.

## CONFLICT OF INTEREST STATEMENT

The authors declare no conflicts or competing interests.

## SOURCES OF FUNDING

This work was supported by funds from the Program for Neuropsychiatric Research, McLean Hospital (BMC) and NIGMS: R35GM134949 (IL).

## CONSENT STATEMENT

All subjects that participated in clinical trials provided informed consent. In the case of incapacity, consent was provided by an authorized representative/health-care proxy. All authors read, edited, and approved the final manuscript and gave consent for publication.

## Notes

### Competing Interest Statement

The authors have declared no competing interest.

### Author Declarations

IRB of Mass General Brigham gave ethical approval for this work.

