## Supplemental figure and Tables for "Specific Lipid Abnormalities Are Inherently Associated with Late-Onset Alzheimer’s Disease"

**Supplementary Fig S1. Purity of mitochondria**

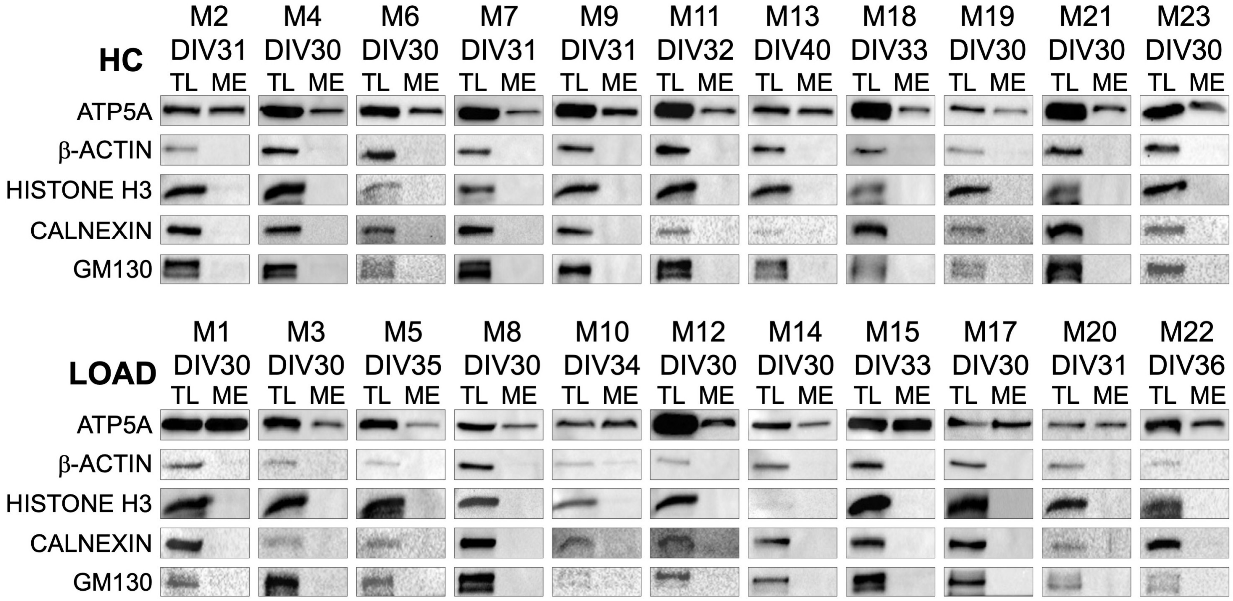


Mitochondrial extracts (ME) and total cell lysates (TL) were analyzed for mitochondrial (ATP5A), cytoplasmic (b-ACTIN), nuclear (HISTONE H3), endoplasmic reticulum (CALNEXIN), or Golgi (GM 130) markers. Samples were coded for blind analysis. Day in vitro (DIV) depicts day of astrocyte differentiation on which cells were lysed and mitochondria were extracted.

**SupplementaryTable S1.  Antibodies**

| Name | Marker | Size (kDA) | Dilution | Hybridization | Catalog number (Provider) | RRID |
|---|---|---|---|---|---|---|
| ATP5A | Mitochondria | 53 | 1:2000 | Overnight at 4 ºC | ab14748 abcam | AB_301447 |
| Histone H3 | Nucleus | 17 | 1:1000 | One hour at RT followed by overnight at 4 ºC | 9715S Cell Signaling Technology, MA, USA | AB_331563 |
| ß-actin | Cytosol | 43 | 1:2000 | Overnight at 4 ºC | sc-47778 Santa-Cruz Biotechnology, TX, USA | AB_626632 |
| Calnexin | Endoplasmic reticulum | 90 | 1:500 | One hour at RT followed by overnight at 4 ºC | sc-23954 Santa-Cruz Biotechnology | AB_626783 |
| GM130 | Golgi | 130 | 1:2000 | One hour at RT followed by overnight at 4 ºC | 12480S Cell Signaling Technology | AB_2797933 |

**RRID: Research Resource Identifiers**

**SupplementaryTable S2.  Lipid Classes**

| Abbreviation | Full Name | Whole Cells | Mitochondria |
|---|---|---|---|
| **CE** | Cholesteryl ester | V | △ |
| **Cer** | Ceramide | V | V |
| **Chol** | Cholesterol | V | V |
| **DAG** | Diacylglycerol | V | V |
| **DiHexCer** | Dihexosyl ceramide | V | △ |
| **GD1** | Ganglioside GD1 | △ | △ |
| **GD2** | Ganglioside GD2 | △ | |
| **GD2-OAc** | Ganglioside GD2 OAc | △ | |
| **GD3** | Ganglioside GD3 | △ | △ |
| **GD3-OAc** | Ganglioside GD3 OAc | △ | |
| **GM1** | Ganglioside GM1 | △ | △ |
| **GM2** | Ganglioside GM2 | △ | △ |
| **GM3** | Ganglioside GM3 | V | V |
| **GT1** | Ganglioside GT1 | △ | |
| **HexCer** | Hexosylceramide | V | V |
| **LPA** | Lyso-phosphatidate | V | |
| **LPC** | Lyso-phosphatidylcholine | V | V |
| **LPC O-** | Lyso-phosphatidylcholine (-ether) | V | △ |
| **LPE** | Lyso-phosphatidylethanolamine | V | V |
| **LPE O-** | lyso-phosphatidylethanolamine (-ether) | V | V |
| **LPG** | Lyso-phosphatidylglycerol | V | |
| **LPI** | Lyso-phosphatidylinositol | V | △ |
| **LPS** | Lyso-phosphatidylserine | V | △ |
| **PA** | Phosphatidic acid | V | V |
| **PC** | Phosphatidylcholine | V | V |
| **PC O-** | Phosphatidylcholine (-ether) | V | V |
| **PE** | Phosphatidylethanolamine | V | V |
| **PE O-** | Phosphatidylethanolamine (-ether) | V | V |
| **PG** | Phosphatidylglycerol | V | V |
| **PI** | Phosphatidylinositol | V | V |
| **PS** | Phosphatidylserine | V | V |
| **SM** | Sphingomyelin | V | V |
| **TAG** | Triacylglycerol | V | △ |

△: detected but filtered out; V: detected and included in the analysis
